# At Least Three Doses of Leading Vaccines Essential for Neutralisation of SARS-CoV-2 Omicron Variant

**DOI:** 10.1101/2022.02.20.22271237

**Authors:** Nagendrakumar B Singanallur, Petrus Jansen van Vuren, Alexander J McAuley, Matthew P Bruce, Michael J Kuiper, Stella M Gwini, Shane Riddell, Sarah Goldie, Trevor W Drew, Kim R Blasdell, Mary Tachedjian, Shruthi Mangalaganesh, Simran Chahal, Leon Caly, Julian D Druce, Jennifer A Juno, Stephen J Kent, Adam K Wheatley, Seshadri S Vasan

**Affiliations:** Commonwealth Scientific and Industrial Research Organisation (CSIRO) Australian Centre for Disease Preparedness (ACDP), Geelong, VIC 3220, Australia; Data61, Commonwealth Scientific and Industrial Research Organisation (CSIRO), Docklands, VIC 3008, Australia; University Hospital Geelong, Barwon Health, Geelong, VIC 3220, Australia; Monash Biomedicine Discovery Institute, Monash University, Clayton, VIC 3168, Australia; Victorian Infectious Diseases Reference Laboratory (VIDRL), The Peter Doherty Institute for Infection and Immunity, Melbourne, VIC 3000, Australia; Department of Microbiology and Immunology, University of Melbourne, The Peter Doherty Institute for Infection and Immunity, Melbourne, VIC 3000, Australia; Department of Health Sciences, University of York, York, YO10 5DD, United Kingdom

**Keywords:** AlphaFold, BA.1.1, biomolecular modelling, COVID-19, neutralising antibody titres, SARS-CoV-2, Spike protein, variants of concern, vaccines, vaccine matching

## Abstract

Plasma samples taken at different time points from donors who received either AstraZeneca (Vaxzevria) or Pfizer (Comirnaty) or Moderna (Spikevax) coronavirus disease-19 (COVID-19) vaccine were assessed in virus neutralization assays against Delta and Omicron variants of concern and a reference isolate (VIC31). With the Pfizer vaccine there was 6-8-fold reduction in 50% neutralizing antibody titres (NT_50_) against Delta and VIC31 at 6 months compared to 2 weeks after the second dose; followed by 25-fold increase at 2 weeks after the third dose. Neutralisation of Omicron was only consistently observed 2 weeks after the third dose, with most samples having titres below the limit of detection at earlier timepoints. Moderna results were similar to Pfizer at 2 weeks after the second dose, while the titres for AstraZeneca samples derived from older donors were 7-fold lower against VIC31 and below the limit of detection against Delta and Omicron. Age and gender were not found to significantly impact our results. These findings indicate that vaccine matching may be needed, and that at least a third dose of these vaccines is necessary to generate sufficient neutralising antibodies against emerging variants of concern, especially Omicron, amidst the challenges of ensuring vaccine equity worldwide.

## Introduction

The novel coronavirus disease-19 (COVID-19) pandemic caused by the severe acute respiratory syndrome coronavirus-2 (SARS-CoV-2) has resulted in over half a billion cases and 6.2 million deaths, and is yet to be brought under control [**1**]. Only 14.2 vaccine doses have been administered per 100 people in ‘low-income countries’; for ‘lower middle income countries’ excluding India the number is 81.6 doses per 100 people [**2**]. Thus, half the world’s population is still receiving the first dose of a COVID-19 vaccine and is very vulnerable [**3**]. Although vaccines can reduce the likelihood of infection, re-infection and disease severity in individuals, and to an extent community transmission, the continuous emergence of SARS-CoV-2 variants of concern (VOC) has frustrated global response efforts and highlighted the importance of continued assessment of vaccine efficacy [**4,5**]. The World Health Organization has declared 5 VOC to date, viz. Alpha (18 December 2020), Beta (18 December 2020), Gamma (11 January 2021), Delta (11 May 2021), Omicron (26 November 2021), of which Delta and Omicron are currently of the greatest concern and two variants of interest (VOI) viz. Lambda (14 June 2021) and Mu (30 August 2021) [**6**].

While studies on T-cell responses will provide a more holistic picture of vaccine-induced efficacy, assessment of neutralising antibody reactivity against emerging VOC, such as Omicron (B.1.1.529, referred to as BA.1 henceforth), can help inform near-term public health response. This is especially important for the vaccines manufactured by Pfizer, Moderna, and AstraZeneca, as one or more of these three are approved in 192 countries (which does not include the People’s Republic of China), representing 77% of the world’s population [**2**]. In this study we have assessed the ability of human plasma samples collected from vaccinated donors at different timepoints to neutralise infectious SARS-CoV-2 virus isolates of the current two most globally-prevalent VOC (Delta and Omicron; particularly the under-studied Omicron BA.1.1 sub-lineage) compared with a reference isolate used in previous studies (VIC31; [**7**]). This adds to the collective evidence base of the efficacy of existing vaccines against the Omicron VOC (of which there are a number of sub-lineages), from the perspective of different experimental protocols (including cell lines) and regional patient and virus samples [**4,5**]. We have further bolstered our interpretation using *in silico* modelling of the respective Spike proteins and outlined our thoughts on future follow-up studies.

## Materials and Methods

### SARS-CoV-2 Stock Generation and Characterisation

Three SARS-CoV-2 isolates, VIC31 (B.1; hCoV-19/Australia/VIC31/2020 containing D614G mutation), Delta (B.1.617.2) variant of concern (hCoV-19/Australia/VIC18440/2021), and Omicron (BA.1.1) variant of concern (hCoV-19/Australia/VIC28585/2021) were provided by the Victorian Infectious Diseases Reference Laboratory (VIDRL; Melbourne, Australia). Virus stocks were propagated and titrated in Vero E6 cells (American Type Culture Collection (ATCC), Manassas, VA, USA) prior to use as previously outlined in [**8**]. Briefly, Vero E6 cells were grown in 150 cm^2^ flasks in Dulbecco’s Modified Eagle Medium (DMEM) containing 10% heat-inactivated foetal bovine serum (FBS), 2mM GlutaMAX supplement, 100U/mL penicillin, and 100 μg/mL streptomycin (all components from ThermoFisher Scientific; Scoresby, VIC, Australia) until >80% confluent. Virus isolates were diluted 1:100 in DMEM (containing 2% FBS, 2mM GlutaMAX supplement, 100U/mL penicillin, and 100 μg/mL streptomycin), and 5 mL was used to inoculate Vero E6 cells for 1 hr at 37°C/5% CO_2_ before additional media was added to the flask. The flasks were incubated at 37°C/5% CO_2_ for 48 h (for VIC31 and Delta) or 72 hr (for Omicron) before supernatant was clarified at 2,000 x *g* for 10 min, and harvested and stored in 1 mL aliquots at -80°C.

Identity of virus stocks were confirmed by next-generation sequencing using a MiniSeq platform (Illumina, Inc; San Diego, CA, USA). In brief, 100 µL cell culture supernatant from infected Vero E6 cells was combined with 300 µL TRIzol reagent (Thermo Fisher Scientific) and RNA was purified using a Direct-zol RNA Miniprep kit (Zymo Research; Irvine, CA, USA). Purified RNA was further concentrated using an RNA Clean-and-Concentrator kit (Zymo Research), followed by quantification on a DeNovix DS-11 FX Fluorometer. RNA was converted to double-stranded cDNA, ligated then isothermally amplified using a QIAseq FX single cell RNA library kit (Qiagen, Hilden, Germany). Fragmentation and dual-index library preparation was conducted with an Illumina DNA Prep, Tagmentation Library Preparation kit. Average library size was determined using a Bioanalyser (Agilent Technologies; San Diego, CA, USA) and quantified with a Qubit 3.0 Fluorometer (Invitrogen; Carlsbad, CA, USA). Denatured libraries were sequenced on an Illumina MiniSeq using a 300-cycle Mid-Output Reagent kit as per the manufacturer’s protocol. Paired-end Fastq reads were trimmed for quality and mapped to the published sequence for the SARS-CoV-2 reference isolate Wuhan-Hu-1 (RefSeq: NC_045512.2) using CLC Genomics Workbench version 21 from which consensus sequences were generated. Stocks were confirmed to be free from contamination by adventitious agents by analysis of reads that did not map to SARS-CoV-2 or cell-derived sequences.

### Human Plasma Samples

Blood samples in EDTA blood collection tubes (BD Biosciences, Australia) were collected from healthy volunteers, aged between 25-70 (both female and male), who received COVID-19 vaccine manufactured by either AstraZeneca (Vaxzevria/AZD1222; University of Oxford-AstraZeneca, Oxford/Cambridge, UK), Pfizer (Comirnaty/BNT162b2; BioNTech-Pfizer, Brooklyn, New York, USA), or Moderna (Spikevax/mRNA-1273; Moderna Inc, Cambridge, MA, USA) (**Table 1**). AstraZeneca vaccine was advised for the age group >60 [**9**]. All groups received the same vaccine (homologous) for first and second doses. Most of the Pfizer vaccine group also received a third dose as homologous booster, while the groups that received AstraZeneca or Moderna did not post-third dose samples available. Plasma was separated and stored at -70°C until use. The study protocols were approved by institutional ethics committees of the Peter Doherty Institute, Melbourne Australia [University of Melbourne Central Human Research Ethics Committee (2021-21198-15398-3)] and the CSIRO (CSIRO Human Research Ethics Committee ID 2021_123_RR). Biosafety protocols for handling the human samples and the infectious agents were approved by the Institutional Biosafety Committee of ACDP, Geelong before infectious work commenced. Blood samples were collected on the day of vaccination (baseline; pre-1^st^ dose) and 2 weeks post second dose (2wk-2^nd^ dose) for all the vaccine groups and after six months of 2^nd^ dose (6mo-2^nd^ dose) and two weeks post third dose (2wk-3^rd^ dose) for the Pfizer group.

**Table 1:** Information on Human Participants, Vaccines, and Blood Collection Schedule.

|  |  | AstraZeneca | Moderna | Pfizer |
| --- | --- | --- | --- | --- |
| Donor |  | n=6 | n=12 | n=15 |
| Sex | Male | n=3 | n=6 | n=5 |
|  | Female | n=3 | n=6 | n=10* |
| Age<br>[Median (Range)] | Male | 58 (57-65) | 41.5 (29-70) | 31 (29-35) |
|  | Female | 57 (31-59) | 38.5 (27-47) | 33.5 (25-57) |
|  | Overall | 57.5 (31-65) | 38.5(27-70) | 33 (25-57) |
| Samples Used | pre-1<sup>st</sup> Dose | n=6 (3M, 3F) | n=12 (6M, 6F) | n=15 (5M, 10F) |
|  | 2w Post-2<sup>nd</sup> Dose | n=6 (3M, 3F) | n=12 (6M, 6F) | n=15 (5M, 10F) |
|  | 6mo Post-2<sup>nd</sup> Dose |  |  | n=15 (5M, 10F) |
|  | 2w Post-3<sup>rd</sup> Dose |  |  | n=14 (4M, 10F) |

\*For pre-1<sup>st</sup> dose and 2w Post-2<sup>nd</sup> dose the samples are gender balanced.

### Live-Virus Neutralisation Assays

Virus neutralization assays (VNT) were carried out using Vero E6 cells as described previously [**8**]. Briefly, each plasma sample for the three vaccines was diluted 1:10 in Dulbecco’s Phosphate Buffered Saline (DPBS; Thermo Fisher Scientific, Waltham, MA, USA) in a deep-well plate, followed by a two-fold serial dilution up to 1:1,280. The dilution series for each plasma sample was dispensed into triplicate columns of a 96-well plate (one plate per isolate), for a total volume of 50 µl per well. For the plasma-containing wells, 50 µl virus diluted in DMEM (Thermo Fisher Scientific, Waltham, MA, USA) containing 2% FBS, 2mM GlutaMAX supplement, 100U/mL penicillin, and 100 μg/mL streptomycin to contain approximately 100 TCID_50_ (checked by back-titration) was added to each well. The plates were incubated at 37°C/5% CO_2_ for 1 h to allow neutralization complexes to form between the antibodies present in the plasma and the virus. A positive control serum was included to confirm the reproducibility of the assay. At the end of the incubation, 100 µl Vero E6 cells (2×10^4^ cells/well) were added to each well and the plates were returned to the incubator for 4 days. Each well was scored for the presence of viral CPE, readily discernible on Day 4 post-infection. NT_50_ neutralization titres calculated using the Spearman–Kärber formula [**10**] and transformed to log_2_ values for analysis. Replicates that did not show neutralisation at 1:10 were scored <1:10 dilution and assigned a value 1:5 for statistical analysis in line with the current best practice [**11**].

### *In silico* Modelling

Molecular models were made of the trimeric Omicron Spike protein from residues 13 to 1160 (omitting the transmembrane domain) consistent with variant BA.1.1 and including glycosylation to visualize structural changes from the original strain. ‘AlphaFold’ [**12**] was used to reconstruct the N-terminal domain due to insertions and deletions. Models were built to include the ‘up’ and ‘down’ conformations of the Receptor Binding Domain (RBD, residues 330-530) as well as the human angiotensin converting enzyme 2 (ACE2) receptor. Building upon our previous work [**13**], models were solvated and simulated using ‘NAMDv2.14’ software [**14**] for over 200 nanoseconds before visual assessment of changes to surface epitopes and receptor binding interfaces. More modelling details included in Supplementary Methods.

### Statistical Analysis

NT_50_ values were analysed and expressed as Log base 2 (Log_2_). The VNT data were grouped based on vaccines, sex (female and male), and age groups (<35 – young; 35-60 – middle and >60 – senior). Both one-way and two-way Analysis of Variance (ANOVA) were used to test the statistical differences between vaccines, sex, age, day post vaccination/booster, variants, and their interactions. If the ANOVA results returned a p-value of <0.05, a *post hoc* test with a Tukey’s range test (Tukey’s Honestly Significant Difference, c.f. Supplementary Tables) was performed to measure the interactions between two variables. Plots were drawn using ‘*ggplot2*’ and modified in Adobe Illustrator for clarity. All statistical procedures used the ‘*car’* and ‘*lme4’* libraries in R [**15**]. Fold changes were calculated as: 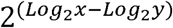.

## Results and Discussion

### Neutralisation of Delta and Omicron Variants of Concern

As neutralising antibody titres are a known correlate of protection for SARS-CoV-2 [**16**], neutralisation assays were performed against SARS-CoV-2 VIC31, Delta, and Omicron using plasma samples collected from human volunteers. **Table 1** shows the median ages for AstraZeneca sample donors are 57.5 compared to 38.5 for Moderna, and 33 for Pfizer vaccine donors respectively, the reasons for which are as follows. The Australian Technical Advisory Group on Immunisation (ATAGI) changed its recommendations regarding the AstraZeneca vaccine following reports of thrombotic thrombocytopenia syndrome (TTS) emerging from vaccinated adults [**17**]. It is worth noting that some individuals younger than 60 have received AstraZeneca vaccine following an assessment by a qualified health processional and with a verbal or written consent as provisioned by the Department Health, Government of Australia [**9**], but there is no brand preference for people aged 60 years and above because the benefits outweigh the risk of TTS in this age group. As a result, samples predominantly originated from young and middle-aged individuals for the Moderna and Pfizer vaccines, and from middle-aged and senior adults for the AstraZeneca vaccine. Given the large age range for the Pfizer vaccine, for this manuscript we classified the age groups in to young (<35 age), middle (35-60) and senior (>60). We chose 35 as the cut-off for middle age as it generally corresponds to parents of teen aged children and is used in metrics such as the Peterson-KFF health System Tracker [**18**] which has identified COVD-19 as the second and third most important cause of death of people who are over and under 35, respectively. In Australia, Moderna vaccine was approved for use relatively recently (9 August 2021), while a third dose of the AstraZeneca is only approved in exceptional circumstances, therefore this study did not have access to third-dose samples for these vaccines.

Calculation of fold changes using the data presented in **Table 2** and **Figure 1**, demonstrate that for the Pfizer vaccine there was an 8.2- and 5.5-fold reduction in NT_50_ titres against VIC31 and Delta respectively 6 months after second dose administration (compared to two weeks), while the average titre against Omicron decreased from an estimated 1:6.3 (only 4/15 samples had quantifiable neutralization) to below assay detection limit for all samples at the same time points, demonstrating that levels of circulating neutralising antibodies decrease in the months following vaccination. Subsequently there was a 25.5- and 24.6-fold increase in titres against VIC31 and Delta respectively two weeks after a third dose administration (compared to 6 month post-second dose), while the average titre against Omicron increased from below assay detection limit to 1:48.5 at the same time points. Because of the aforementioned limitations, calculation of such fold changes for AstraZeneca and Moderna were not possible. However, it is worthwhile to compare their neutralisation titres two weeks post-second dose relative to the Pfizer vaccine samples. Across all three variants, we did not find any significant difference between Pfizer and Moderna vaccines by 2^nd^ week post second dose of the vaccines (**Supplementary Table S1**). Interestingly, the neutralisation titres against VIC31 were generally lower than have been reported in other studies with equivalent samples and virus isolates. This is likely a reflection of the protocol and cells used in this study, and the age groups/populations from which the samples were collected [**4**].

**Table 2:** Sex-Based Mean and Standard Deviation for VNT Titres Against SARS-CoV-2 Variants for Human Subjects Vaccinated with Different COVID-19 Vaccines.

| Vaccine | Sex | Estimates | VIC31 |  |  |  | Delta |  |  |  | Omicron |  |  |  |
| --- | --- | --- | --- | --- | --- | --- | --- | --- | --- | --- | --- | --- | --- | --- |
|  |  |  | 1 | 2 | 3 | 4 | 1 | 2 | 3 | 4 | 1 | 2 | 3 | 4 |
| AstraZeneca | Female | Mean | 2.32 | 3.46 | X | X | 2.32 | 2.32 | X | X | 2.32 | 2.32 | X | X |
|  |  | SD | 0.00 | 0.00 | X | X | 0.00 | 0.00 | X | X | 0.00 | 0.00 | X | X |
|  | Male | Mean | 2.32 | 5.04 | X | X | 2.32 | 2.82 | X | X | 2.32 | 2.32 | X | X |
|  |  | SD | 0.00 | 1.01 | X | X | 0.00 | 0.86 | X | X | 0.00 | 0.00 | X | X |
|  | All | Mean | 2.32 | 4.25 | X | X | 2.32 | 2.57 | X | X | 2.32 | 2.32 | X | X |
|  |  | SD | 0.00 | 1.08 | X | X | 0.00 | 0.61 | X | X | 0.00 | 0.00 | X | X |
| Moderna | Female | Mean | 2.32 | 7.16 | X | X | 2.32 | 5.99 | X | X | 2.32 | 2.57 | X | X |
|  |  | SD | 0.00 | 1.11 | X | X | 0.00 | 0.76 | X | X | 0.00 | 0.61 | X | X |
|  | Male | Mean | 2.32 | 6.93 | X | X | 2.32 | 5.05 | X | X | 2.32 | 2.82 | X | X |
|  |  | SD | 0.00 | 1.17 | X | X | 0.00 | 1.28 | X | X | 0.00 | 0.80 | X | X |
|  | All | Mean | 2.32 | 7.04 | X | X | 2.32 | 5.52 | X | X | 2.32 | 2.69 | X | X |
|  |  | SD | 0.00 | 1.09 | X | X | 0.00 | 1.12 | X | X | 0.00 | 0.69 | X | X |
| Pfizer | Female | Mean | 2.32 | 7.22 | 3.75 | 8.93 | 2.32 | 6.36 | 3.37 | 7.39 | 2.32 | 2.73 | 2.32 | 5.95 |
|  |  | SD | 0.00 | 1.08 | 0.66 | 1.07 | 0.00 | 1.04 | 0.79 | 0.82 | 0.00 | 0.67 | 0.00 | 1.22 |
|  | Male | Mean | 2.32 | 6.76 | 4.61 | 9.32 | 2.32 | 5.76 | 4.34 | 8.82 | 2.32 | 2.49 | 2.32 | 6.07 |
|  |  | SD | 0.00 | 0.92 | 0.71 | 0.64 | 0.00 | 1.06 | 0.97 | 1.09 | 0.00 | 0.38 | 0.00 | 1.43 |
|  | All | Mean | 2.32 | 7.07 | 4.04 | 9.04 | 2.32 | 6.16 | 3.70 | 8.23 | 2.32 | 2.65 | 2.32 | 5.99 |
|  |  | SD | 0.00 | 1.02 | 0.78 | 0.96 | 0.00 | 1.05 | 0.95 | 0.94 | 0.00 | 0.59 | 0.00 | 1.23 |

All VNT titres expressed as Log2 values. Values indicated as 2.32 represent titres below the assay detection limit. SD = Standard Deviation of Mean; 1 = pre-1<sup>st</sup> dose; 2 = 2wk Post-2<sup>nd</sup> dose; 3 = 6mo Post-2<sup>nd</sup> dose and 4 = 2wk Post-3<sup>rd</sup> dose. X = sample not available for analysis

**Figure 1:**
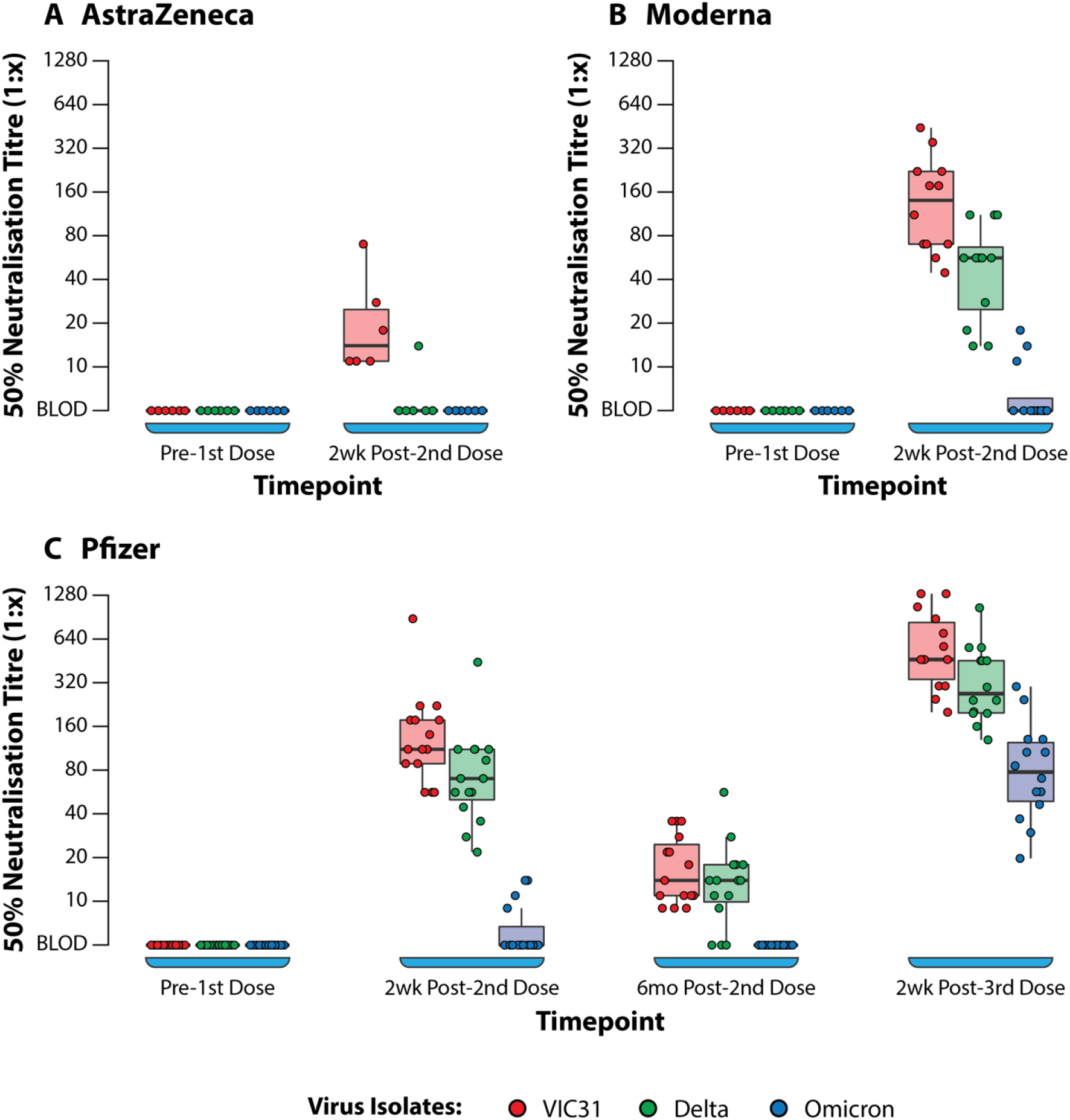
Box Plots Showing 50% Neutralisation Titres (NT_50_) Against SARS-CoV-2 Variants of Concern with Plasma from Human Volunteers. Neutralisation of VIC31, Delta, and Omicron by human donor samples collected at different timepoints following vaccination with AstraZeneca (A), Moderna (B), or Pfizer (C) COVID-19 vaccines. The bold horizonal line represents median titre, with the 1st and 3rd quartiles represented by the box. Statistical analysis is provided in Supplementary Table S3.

ANOVA (one-way and two-way) was used to investigate the interactions of age, sex, vaccine, and day post vaccination/booster with neutralising antibody titres to the three different virus variants. One-way and two-way ANOVA analyses showed that, for all vaccines (Pfizer, Moderna and AstraZeneca), day post vaccination/booster and virus variant were the main contributors to differences in neutralising antibody responses, while age and sex did not have significant effects.

Based on ANOVA analysis there was no significant difference in neutralising titres based on age or sex for any of the vaccines (**Supplementary Tables S2 & S3; Figure 2A-C**). The titres were dependent on the vaccine, day post-vaccination and variant (p<0.001) and to an extent the age where antibody titres in young showed statistically significant difference when compared to the middle and senior age groups (p=0.022). The antibody titres against Omicron have a negative slope and indicating that there is poor neutralisation with the variant.

**Figure 2:**
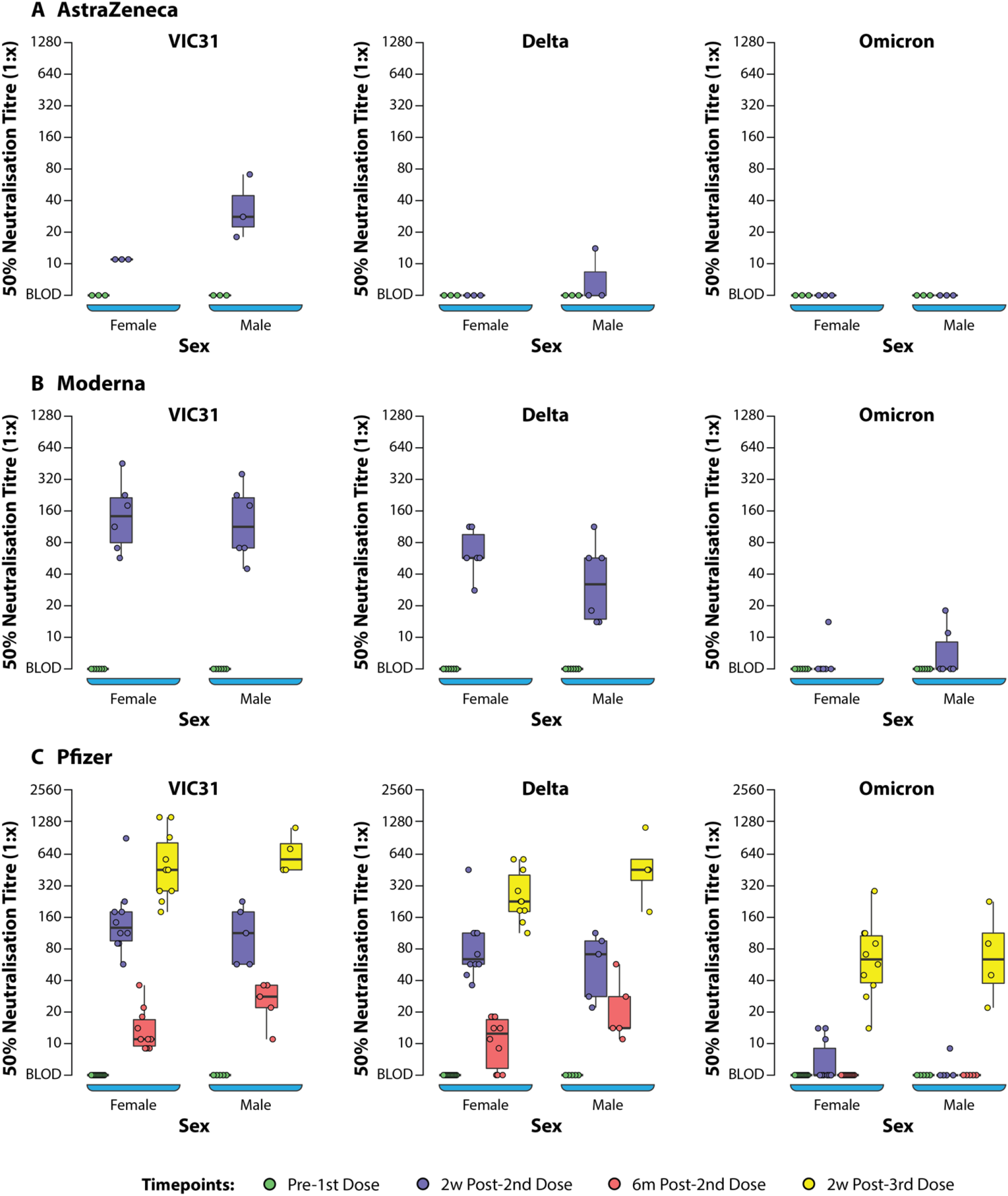
Box Plots Showing 50% Neutralization Titres (NT_50_) Against SARS-CoV-2 Variants of Concern Assessed by Sex of Donors. Plasma samples from human volunteers vaccinated with either AstraZeneca (A), Moderna (B), or Pfizer (C) vaccines were tested in neutralisation assays against VIC31 (i), Delta (ii), and Omicron (iii), with titres assessed with respect to sex. The bold horizonal line represents median titre, with the 1st and 3rd quartiles represented by the box. Statistical analysis is provided in Supplementary Table S3.

**Figure 3:**
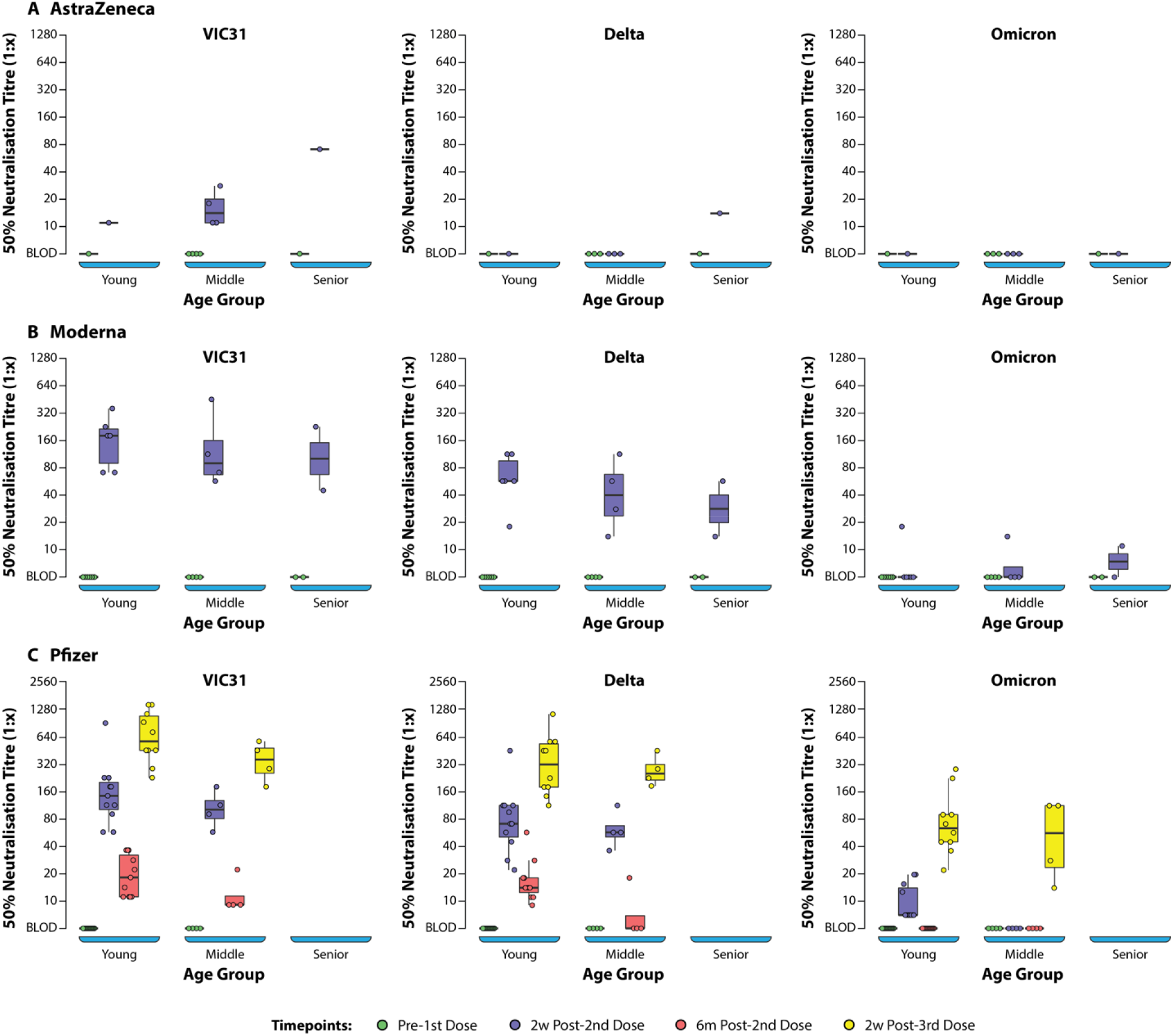
Box Plots Showing NT_50_ Values Against SARS-CoV-2 Variants of Concern Assessed by Age Group. Plasma samples from human volunteers vaccinated with either AstraZeneca (A), Moderna (B), or Pfizer (C) vaccines were tested in neutralisation assays against VIC31 (i), Delta (ii), and Omicron (iii), with titres assessed with respect to age group. The bold horizonal line represents median titre, with the 1st and 3rd quartiles represented by the box. Statistical analysis is provided in Supplementary Table S3.

### Importance of Third Dose for Neutralisation of Omicron

In line with recent findings in similar studies by other groups [**19,20**], Omicron titres were significantly lower than VIC31-D614G and Delta titres at all time points and regardless of vaccine, highlighting the concern of poor protection to Omicron following vaccination by any of the three leading vaccines. One shortcoming of our data is the lack of later samples and post-boost (third vaccination) from individuals vaccinated with AstraZeneca and Moderna vaccines. However, the data available from Pfizer vaccinated individuals at 6 months after receiving their second dose indicate a significant waning of immunity to all three variants over this period. Neutralising antibody levels to Omicron were low or undetectable in most individuals two weeks after receiving two Pfizer doses, and below the assay limit of detection in all samples tested at 6 months after the second dose. However, at two weeks after the booster vaccination (third Pfizer dose), a significant booster effect in neutralising titres was noted to all three variants. This highlights the importance of the need for a booster (third) vaccine dose for ongoing protection against D614G and Delta variants, but more importantly that two doses are insufficient to stimulate significant neutralising protection against Omicron BA.1, therefore making the third dose essential. The cause of better protection against Omicron BA.1 following three doses of vaccine incorporating Spike protein lacking Omicron specific mutations, is likely due to higher overall neutralising titres due to a booster effect despite neutralising antibodies to certain epitopes being circumvented.

### Modelling and Analyses using *in silico* methods

In order to identify a mechanistic explanation for the observed reduction in neutralisation of Omicron compared to other variants, we investigated the amino acid mutations present in Omicron. Consistent with other studies, there are significantly more mutations in the Spike protein of the Omicron variant shown in **Figure 4A**, (37 mutations in the BA.1 variant, 32 in the BA.2 variant including insertions and deletions), compared to the Delta variant (10 mutations), while only one in the VIC31 strain (D614G). The Omicron mutations contribute to its ability to circumvent vaccine or infection induced neutralising antibody responses [**19,20**]. Three of these mutations (G142D, T478K and D614G) are common to Delta and Omicron, however we have previously shown experimentally and through *in silico* modelling that D614G has no significant impact on vaccines [**7**]. In the receptor binding domain (RBD) of the Spike protein alone, which is most immunologically exposed in its ‘up’ configuration and critical for ACE2 binding, there are 15 and 17 mutations in the Omicron BA.1 and BA.2 variants compared to only 2 in Delta as shown in **Figures 4A & C**. Many of the observed Omicron mutations by our analysis appear to be immune evasive, as they do not appear to be at the ACE2 binding interface and represent a significant change in the residue characteristics; in the RBD, these include N440K, K417N, S477N, T478K, and E484A.

**Figure 4:**
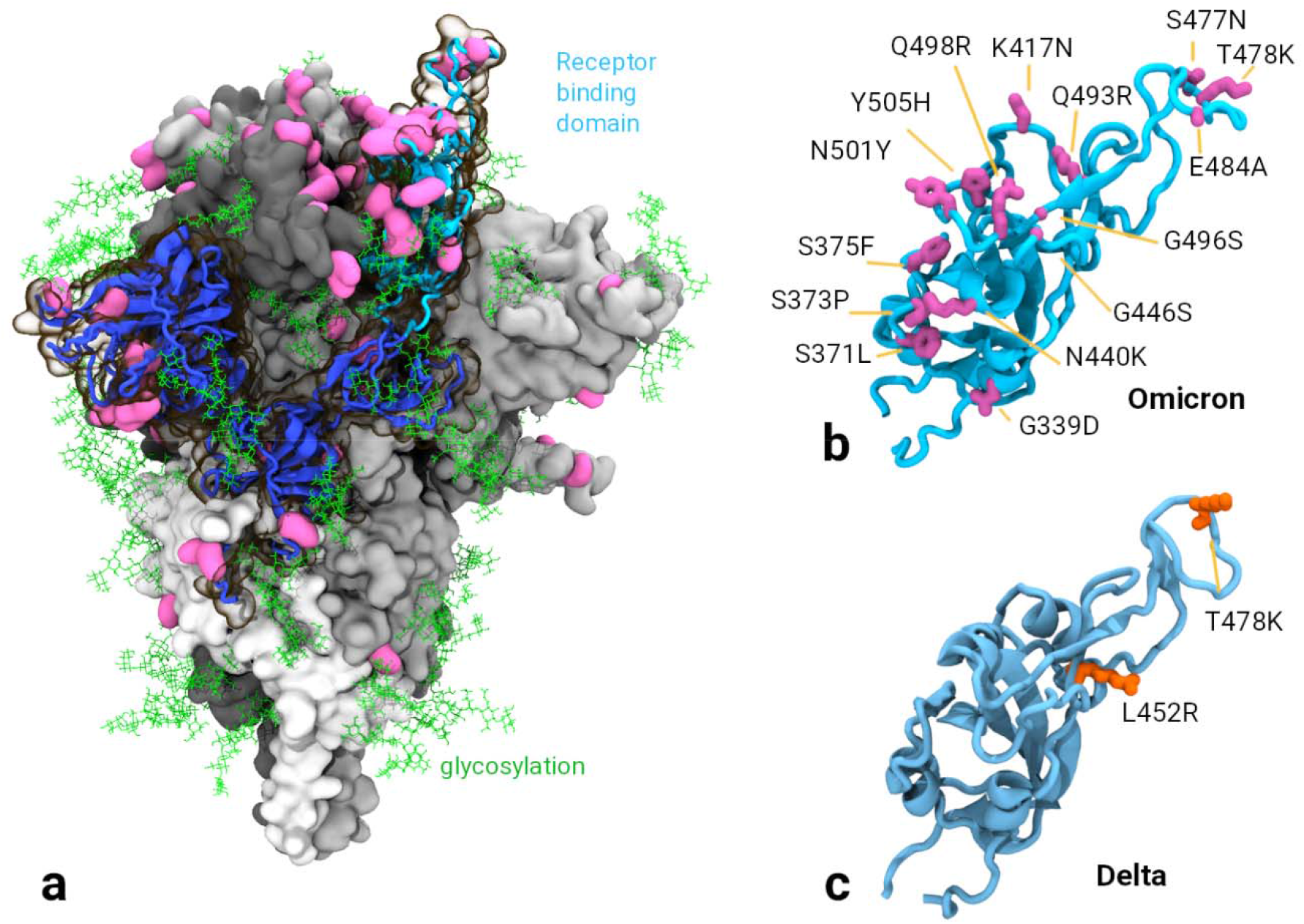
Structure of SARS-CoV-2 Spike Protein and Mutations of Interest. **A**. The Omicron spike protein with mutations shown in pink. A single S1 domain is highlighted in blue with the receptor binding domain (RBD) in the ‘up’ configuration is highlighted in cyan. Glycosylation is shown as green lines. **B**. & **C**. Comparison of the RBD in the Omicron and Delta variants, 15 mutations are observed in Omicron while only 2 in the Delta variant.

Interestingly our isolate VIC28585 belongs to the BA.1.1 sub-lineage as it also contains the R346K spike mutation, which possibly further decreases antibody neutralization based on the reduced binding of class 2 antibodies in theoretical studies with the Mu variant of interest [**21**]. Next-generation sequencing results also indicated the presence of additional Spike mutations S373P and S375F in our VIC28585 isolate at frequency exceeding 99.8%; these two mutations with Grantham scores of 74 (moderately conservative change) and 155 (radical change) respectively are near the S371L Spike mutation characteristic to BA.1 which has a Grantham score of 145 (moderately radical change; *n*.*b*. these three mutations are also present in EPI_ISL_7869197 mentioned below). Thus, we have investigated the neutralising responses to Omicron BA.1.1 in individuals vaccinated with three leading COVID-19 vaccines and compared these neutralising titres to those against an early SARS-CoV-2 isolate with the D614G mutation (VIC31), and the Delta variant (B.1.617.2) that subsequently became the dominant variant worldwide prior to Omicron’s emergence. For example, on 11 December 2021 the ratio of Delta : BA.1 : BA.1.1 : BA.2 : Other in USA (where extensive data is available from the CDC; [**22**]) was 92.5 : 5.2 : 2.2 : 0 : 0.2, and this steadily changed over two months so that on 12 February 2021 the ratio was 0 : 22.9 : 73.2 : 3.9 : 0. This shows that BA.1.1, used in our study, could emerge as the dominant variant although we need to wait for more comprehensive data from around the world – as of 16 February 2022, 719,364 sequences of BA.1 were reported from 139 countries, while 410,015 sequences of BA.1.1 were reported from 127 countries.

Other Omicron mutations are likely to confer increased binding affinity to ACE2 by increasing molecular interactions such as salt bridges and pi-stacking [**12**]; these mutations include Q493R and Q498R (increasing salt bridge interactions with ACE2’s D38 and E35) and N501Y (also seen in Alpha, Beta and Gamma variants, pi-stacking with ACE2’s Y41, shown in **Supplementary Figure S1**. A cluster of mutations S371L, S373P and S375F, interestingly appear co-located with Y505H in adjacent RBD domains in the ‘down’ position. These serine mutations, as well as significantly altering epitope presentation of the RBD domain by creating a more rigid structural motif, may allow H505 to interact with the adjacent F375, potentially modulating ‘up’/’down’ conformations via the ionizable H505, (pK_a_ of 6.0). This structural change may coincide with the observed Omicron preference of infection to the bronchus, where pH is usually lower at 5.5 to 6.5, compared to the lung [**23,24**] (**Supplementary Figure S2**).

With the large number of mutations, it is unsurprising that the Omicron variant displays the observed antibody evasion and replicative advantage over earlier variants as seen in other studies and experimentally observed in this study [**3,25**]. Our biomolecular modelling shows sufficient distinction between BA.1 and BA.2, therefore future studies should experimentally compare the two.

### Public Health Implications and Future Studies

The data we present here highlight the importance of booster vaccination for adequate protection against newly emerging variants such as Omicron BA.1.1, particularly the observation that two doses of the three leading vaccines are not sufficient for neutralisation of Omicron, while a third dose of Pfizer resulted in detectable neutralisation titres in all donor samples. Further studies are needed to perform similar analyses of neutralising titres in individuals receiving “heterologous” booster doses, e.g., initial AstraZeneca vaccination followed by Moderna or Pfizer boost, or Pfizer-Moderna / Moderna-Pfizer.

In the early days of the pandemic, the scientific prediction was that SARS-CoV-2 will have a mutation rate similar to SARS-CoV (0.8-2.4×10^−3^) thanks to the 3′-to-5′ exoribonuclease ‘proofreading mechanism’ in coronaviruses [**26**], which was thought to result in a reduction in the mutation rate (∼10^−6^ per site per cycle) compared to influenza (∼3×10^−5^ per site per cycle) [**27,28**].

Therefore, it was predicted that vaccine matching may not be required as frequently as seasonal influenza (once a year each for Northern and Southern hemispheres), although this was hard to say definitively because SARS-CoV-2’s genetic drift was uncertain with respect to the long-lasting efficacy of vaccine candidates then under development [**29**]. Early studies, for instance between 1 February to 1 May 2020 seemed to confirm the mutation rate prediction with a median estimate of 1.12×10^−3^ per site-year ([**30**]; we believe their upper 95% confidence interval should read 1.85×10^−3^).

Two years on, we have increasing evidence, including from this work, that the primary immunization schedule should consist of three rather than two doses, and indeed many countries are redefining ‘fully vaccinated’ status as three doses of these vaccines [**31**]. It is early to speculate on annual boosters beyond the third dose, although Israel has started to expand its fourth dose administration to groups beyond the immunocompromised [**32,33**]. These developments make it all the more important to ensure vaccine equity across the world, so everyone is adequately protected and highlight the potential need for vaccine matching to increase protection against infection. Vaccines using Spike as the primary antigen should consider including mutations common to BA.1, BA.1.1, and BA.2; development of vaccines with additional antigens, other than Spike, targeting more conserved viral factors should also be explored. This view is further supported by a recent preprint currently under peer review suggesting that, although a fourth dose restored anti-Omicron BA.1 (EPI_ISL_7869197) neutralising antibody titres to a level observed post-third dose, breakthrough Omicron infections were common with high virus titres, albeit with very mild clinical signs [**33,34**].

In this context, it is important to explore vaccines that can better withstand mutations to the Spike protein better, and ideally have less dependence on cold chain. In addition to repeating this work with Omicron BA.2, it is also important to explore the benefits of T-cell immunity and heterologous boosting (which are beyond the scope of this paper; but see for instance these studies [**35-38**]). Another aspect worth exploring is intranasal administration of selected vaccines. In the earliest study (conducted from March 2020) looking at intranasal administration of the AstraZeneca vaccine [**39**], we found that although neutralising antibody titres were 2-fold lower in ferrets receiving two doses of this vaccine intranasally compared to the intramuscular route, the former was more protected using virus shedding as an infection metric [**40**]. These findings were subsequently confirmed in hamsters and non-human primates [**41**], and also reportedly led to a small clinical trial in 30 humans [**42**]. We feel that the intranasal route, and the role of mucosal immunity, needs to be explored more comprehensively given that the AstraZeneca vaccine has the widest reach (183 countries worldwide), with over 2.5 billion doses administered [**43**].

## Data Availability

The original contributions presented in the study are included in the article/Supplementary Material. Further inquiries can be directed to the corresponding author.

## Ethics Statement

The study protocols were approved by institutional ethics committees of the Peter Doherty Institute, Melbourne Australia [University of Melbourne Central Human Research Ethics Committee (2021-21198-15398-3)] and the CSIRO (CSIRO Human Research Ethics Committee ID 2021_123_RR). The reference serum for the neutralisation assay was derived from an animal study reviewed and approved by CSIRO ACDP Animal Ethics Committee (AEC 2004).

## Author Contributions

NBS designed the study with PJvV, AJM, SMG and SSV. PJvV and AJM performed the experiments with NBS, MPB, SR and SG. NBS and SMG analysed the experimental data and provided statistical interpretations, with inputs from PJvV, AJM, TWD and SSV. MJK performed biomolecular modelling and provided structural interpretations. LC and JDD isolated the virus variants, which were sequenced by KRB and MT. JAJ, SJK and AKW provided the human plasma samples and obtained the necessary funding for the same. SM, SC and SSV analysed the health economic implications. NBS, PJvV, AJM, MJK, SM, SC and SSV co-wrote the manuscript, and all authors contributed to revisions and creation of the final version. SV conceived the conceived the project and obtained funding.

## Funding

This work was supported by funding (Principal Investigator: S.S.V.) from the CSIRO’s Precision Health & Responsible Innovation Future Science Platforms, National Health and Medical Research Council (grant MRF2009092), and United States Food and Drug Administration (FDA) Medical Countermeasures Initiative (contract 75F40121C00144). J.A.J., S.J.K. and A.K.W. are grateful to the National Health and Medical Research Council (for grants GNT2002073, MRF2005544, GNT1149990, fellowships and an investigator grant). We also thank the Victorian Government, especially the Victorian Department of Health and Human Services, the major funder of the Victorian Infectious Diseases Reference Laboratory and Barwon Health. The article reflects the views of the authors and does not represent the views or policies of the funding agencies including the FDA.

## Conflict of Interest

All claims expressed in this article are solely those of the authors and do not necessarily represent those of their affiliated organizations, or those of the publisher, the editors and the reviewers. Any product that may be evaluated in this article, or claim that may be made by its manufacturer, is not guaranteed or endorsed by the publisher.

## Acknowledgments

The authors are grateful for support from Professor Sharon Lewin, Professor Deborah Williamson and colleagues at the Peter Doherty Institute for Infection and Immunity (https://www.grid.ac/institutes/grid.483778.7); the CSIRO eHealth programme (particularly Drs Laurence Wilson and Carol Lee); and the CSIRO’s Australian Centre for Disease Preparedness (ACDP; https://www.grid.ac/institutes/grid.413322.5), especially Dr Daniel Layton and Kristen McAuley. We thank Dr Amy Shurtleff of the Coalition for Epidemic Preparedness Innovations and the University of Oxford’s Professors Teresa Lambe and Dame Sarah Gilbert for access to positive control ferret sera from their study (CSIRO ACDP Animal Ethics Committee Approval Reference: AEC 2004).

## Supplementary Material

### Supplementary Tables S1 to S3

**Supplementary Table S1:** Comparison for NT50 values on 2^nd^-2wk.

| Comparison | Variant | Estimate<br>(‘adjusted’ p-value) |
| --- | --- | --- |
| Pfizer vs Moderna | VIC31 | p > 0.100 |
| Pfizer vs Moderna | Omicron | p > 0.100 |
| Pfizer vs Moderna | Delta | p > 0.100 |

**Supplementary Table S2:** One-way ANOVA results for vaccines, variants, sex, age and day post vaccination/boosters.

| Vaccine | Variable | F-value | p-value | Tukey’s HSD post hoc Comparison<br>(‘adjusted’ p-value) |
| --- | --- | --- | --- | --- |
| Pfizer | Sex | 0.002 | >0.100 | No significant difference |
|  | Age | 0.667 | >0.100 | No significant difference |
|  | Day post vaccination/booster | 119.9 | <0.0001 | Pre 1<sup>st</sup> Dose – 2wk Post-2<sup>nd</sup> Dose p < 0.0001<br>Pre 1<sup>st</sup> Dose – 6mo Post-2<sup>nd</sup> Dose p < 0.01<br>Pre 1<sup>st</sup> Dose – 2wk Post-3<sup>rd</sup> Dose p < 0.0001<br>2wk Post 2<sup>nd</sup> Dose – 6mo Post-2<sup>nd</sup> Dose p < 0.0001<br>2wk Post 2<sup>nd</sup> Dose – 2wk Post-3<sup>rd</sup> Dose p < 0.0001<br>6mo Post 2<sup>nd</sup> Dose – 2wk Post-3<sup>rd</sup> Dose p < 0.0001 |
|  | Variant | 15.86 | <0.0001 | Delta – Omicron p < 0.0001<br>VIC31 – Omicron p < 0.0001 |
| Moderna | Sex | 0.104 | >0.100 | No significant difference |
|  | Age | 0.074 | >0.100 | No significant difference |
|  | Day post vaccination/booster | 64.56 | <0.0001 | Pre 1<sup>st</sup> Dose – 2wk Post-2<sup>nd</sup> Dose p < 0.0001 |
|  | Variant | 8.839 | <0.001 | Delta – Omicron p < 0.05<br>VIC31 – Omicron p < 0.001 |
| AstraZeneca | Sex | 1.497 | >0.100 | No significant difference |
|  | Age | 1.401 | >0.100 | No significant difference |
|  | Day post vaccination/booster | 7.722 | <0.01 | Pre 1<sup>st</sup> Dose – 2wk Post-2<sup>nd</sup> Dose p < 0.01 |
|  | Variant | 5.744 | <0.01 | VIC31 – Delta p < 0.05<br>VIC31 – Omicron p < 0.05 |

**Supplementary Table S3:**
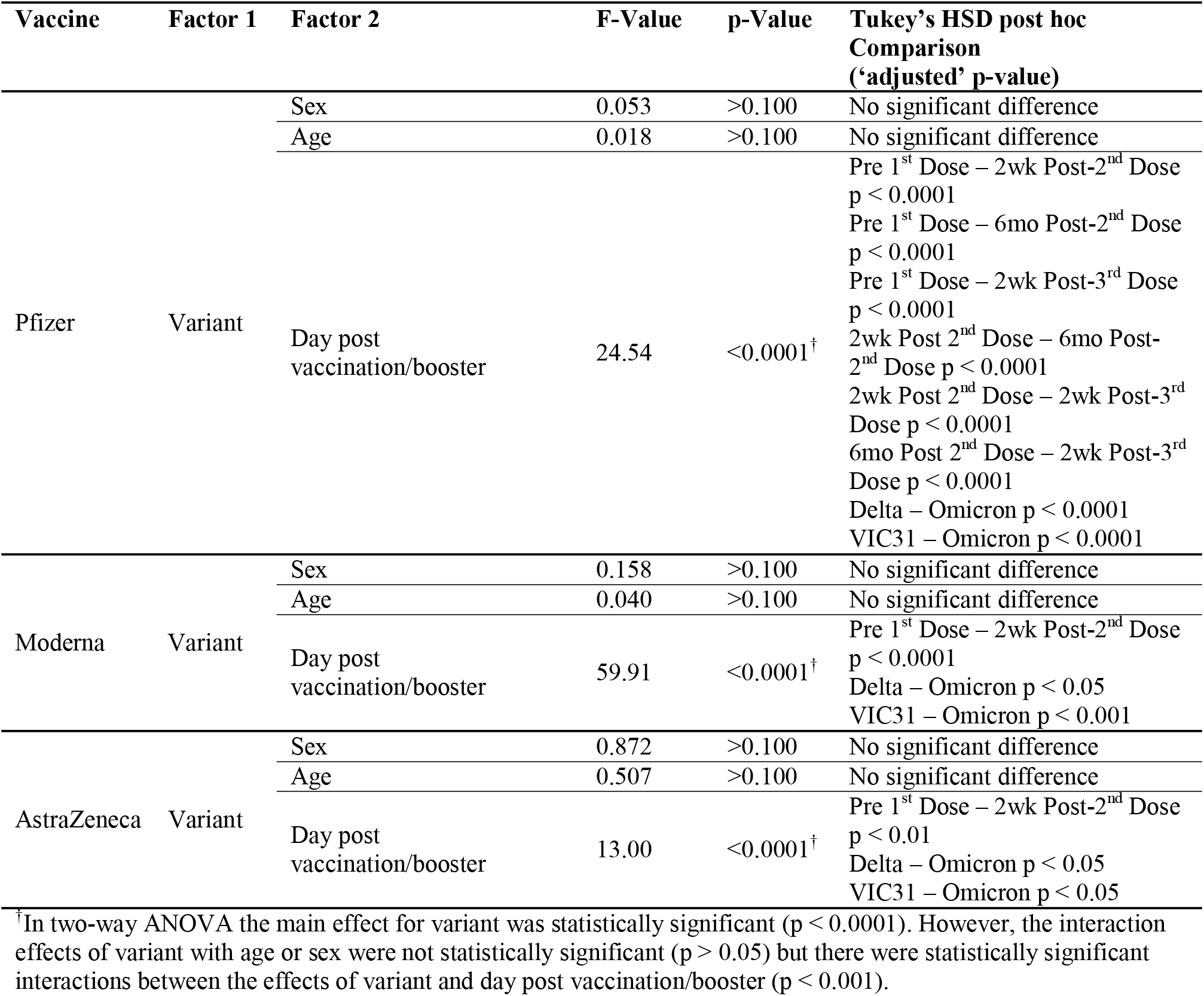
Two-way ANOVA results of interactions for vaccines, variants, sex, age and day post vaccination/boosters.

| Vaccine | Factor 1 | Factor 2 | F-Value | p-Value | Tukey's HSD post hoc Comparison ('adjusted' p-value) |
| --- | --- | --- | --- | --- | --- |
| Pfizer | Variant | Sex | 0.053 | >0.100 | No significant difference |
|  |  | Age | 0.018 | >0.100 | No significant difference |
|  |  | Day post vaccination/booster | 24.54 | <0.0001<sup>†</sup> | Pre 1<sup>st</sup> Dose – 2wk Post-2<sup>nd</sup> Dose p < 0.0001 |
|  |  |  |  |  | Pre 1<sup>st</sup> Dose – 6mo Post-2<sup>nd</sup> Dose p < 0.0001 |
|  |  |  |  |  | Pre 1<sup>st</sup> Dose – 2wk Post-3<sup>rd</sup> Dose p < 0.0001 |
|  |  |  |  |  | 2wk Post 2<sup>nd</sup> Dose – 6mo Post-2<sup>nd</sup> Dose p < 0.0001 |
|  |  |  |  |  | 2wk Post 2<sup>nd</sup> Dose – 2wk Post-3<sup>rd</sup> Dose p < 0.0001 |
|  |  |  |  |  | 6mo Post 2<sup>nd</sup> Dose – 2wk Post-3<sup>rd</sup> Dose p < 0.0001 |
|  |  |  |  |  | Delta – Omicron p < 0.0001 |
|  |  |  |  |  | VIC31 – Omicron p < 0.0001 |
| Moderna | Variant | Sex | 0.158 | >0.100 | No significant difference |
|  |  | Age | 0.040 | >0.100 | No significant difference |
|  |  | Day post vaccination/booster | 59.91 | <0.0001<sup>†</sup> | Pre 1<sup>st</sup> Dose – 2wk Post-2<sup>nd</sup> Dose p < 0.0001 |
|  |  |  |  |  | Delta – Omicron p < 0.05<br>VIC31 – Omicron p < 0.001 |
| AstraZeneca | Variant | Sex | 0.872 | >0.100 | No significant difference |
|  |  | Age | 0.507 | >0.100 | No significant difference |
|  |  | Day post vaccination/booster | 13.00 | <0.0001<sup>†</sup> | Pre 1<sup>st</sup> Dose – 2wk Post-2<sup>nd</sup> Dose p < 0.01 |
|  |  |  |  |  | Delta – Omicron p < 0.05<br>VIC31 – Omicron p < 0.05 |

<sup>†</sup>In two-way ANOVA the main effect for variant was statistically significant (p < 0.0001). However, the interaction effects of variant with age or sex were not statistically significant (p > 0.05) but there were statistically significant interactions between the effects of variant and day post vaccination/booster (p < 0.001).

## Supplementary Methods

### Statistical analysis using linear mixed models

Linear Mixed Models (*lm* and *lmer*) analyses were used to investigate the interactions of age, sex, vaccine, and day post vaccination/booster with neutralising antibody titres to the three different virus variants using’*lme4’* library in R [**15**]. To avoid, over fitting of data, we are not showing the results of *lm* and *lme* analyses. One-way and two-way ANOVA analyses showed that, for all vaccines (Pfizer, Moderna and AstraZeneca), day post vaccination/booster and virus variant were the main contributors to differences in neutralising antibody responses, while age and sex did not have significant effects.

Linear Mixed Model analysis of the variance between variables (results not shown) confirmed the ANOVA analyses which showed that day post vaccination/booster and virus variant are the major contributors to titre differences, but further shows that there was a significant difference in neutralising titres elicited by the three different vaccines, mostly due to poor responses to the AstraZeneca vaccine. This analysis model further suggests that younger individuals (aged less than 35 years) are likely to have a stronger neutralising antibody response than older individuals, but we concede that this finding might be confounded by the distribution of the ages within the vaccine groups. This might also explain the poorer responses in the AstraZeneca group because of the vaccine advice in Australia (AstraZeneca mostly used in the older age group).

### Modelling using *in silico* methods

Molecular simulations were performed using NAMD2.14 with CHARM36m forcefield [**44**] employing a TIP3 water model. The Spike model was based on the pdb structure 6VSB3 [**45**], built with additions segments predicted with AlphaFold. The Spike protein only included residues 13 to 1160 (omitting transmembrane domain, to reduce the simulation size). Glycosylation of the Spike protein was manually constructed using Visual Molecular Dynamics (VMD) guided by glycan analysis [**46**]. Omicron models were constructed in two conformations, all RBD ‘down’, and 2 RBD ‘down’ one RBD ‘up’. Both models included an ACE2 domain in the expected binding position.

Simulations were run with Periodic Boundary Conditions ‘PBCs’ using the NPT ensemble at 310K and 1 bar pressure employing Langevin dynamics. The PBCs were constant in the XY dimensions. Long-range Coulomb forces were computed with the Particle Mesh Ewald method with a grid spacing of 1 Å. 2 fs timesteps were used with non-bonded interactions calculated every 2 fs and full electrostatics every 4 fs while hydrogens were constrained with the ‘SHAKE’ algorithm. The cut-off distance was 12 Å with a switching distance of 10 Å and a pair-list distance of 14 Å. Pressure was controlled to 1 atmosphere using the NoséHoover Langevin piston method employing a piston period of 100 fs and a piston decay of 50 fs. Trajectory frames were captured every 100 ps. Simulations were performed for at least 200 nanoseconds. Trajectories were visualized and analysed using VMD [**47**].

## Supplementary Figures

**Supplementary Figure S1.**
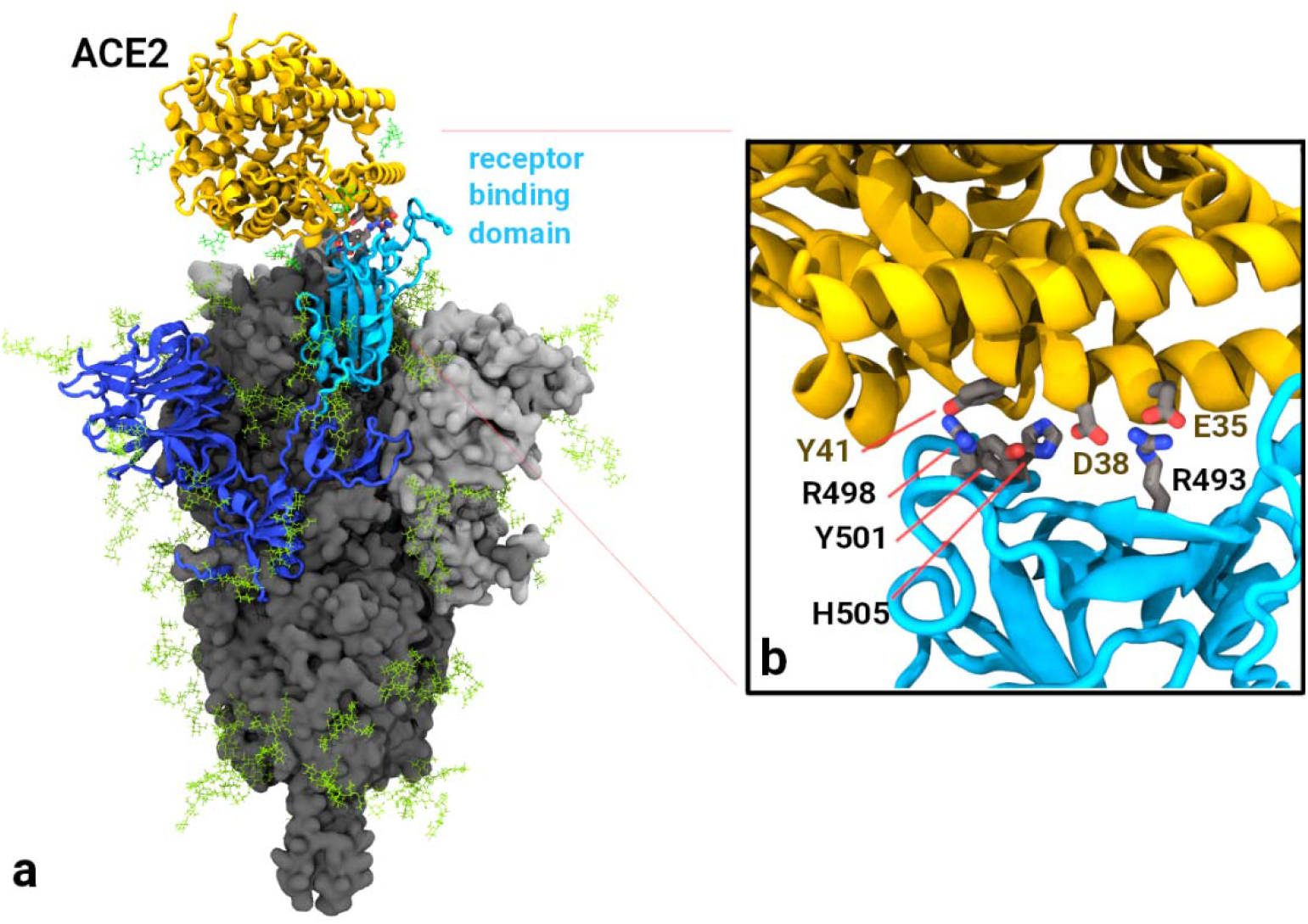
a) Omicron spike protein binding to ACE2 domain (shown in yellow). S1 domain shown in blues, with the receptor binding domain shown in cyan. Glycosylation is shown as green sticks. b) close up of the ACE2 binding interface of Omicron spike RBD showing the interaction of contacting residues.

**Supplementary Figure S2.**
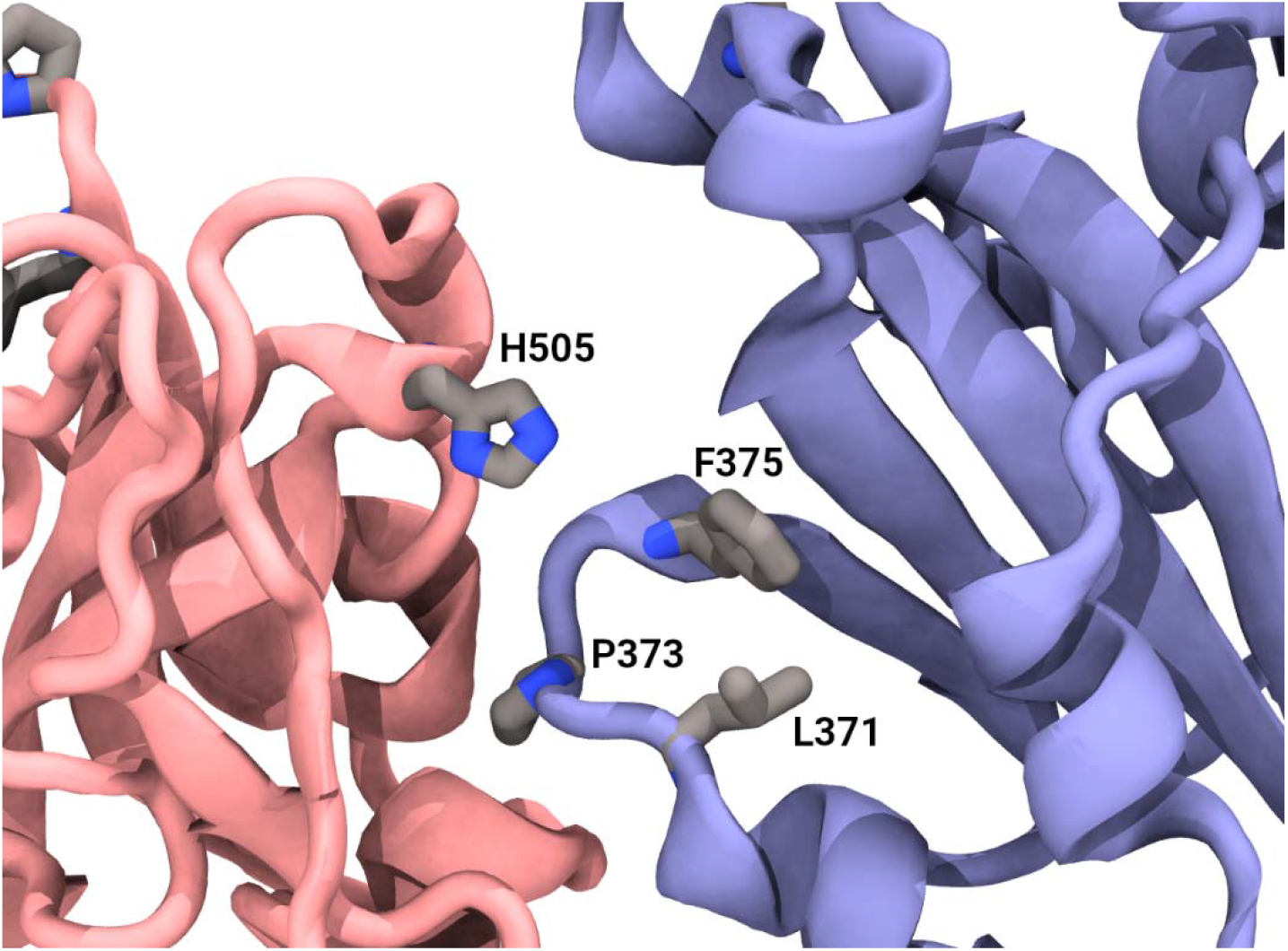
Close up of Omicron mutations in adjacent receptor binding domains (pink and blue) in ‘down’ position showing the close proximity of residues L371, P373 and F375 on the blue domain to the adjacent H505. This complementary arrangement may have influence on the RBD ‘up’/’down’ transitions by pH dependant ionization of H505.

## Notes

### Competing Interest Statement

The authors have declared no competing interest.

### Funding Statement

This work was supported by funding (Principal Investigator: S.S.V.) from the CSIRO Precision Health & Responsible Innovation Future Science Platforms, National Health and Medical Research Council (grant MRF2009092), and United States Food and Drug Administration (FDA) Medical Countermeasures Initiative (contract 75F40121C00144). J.A.J., S.J.K. and A.K.W. are grateful to the National Health and Medical Research Council (for grants GNT2002073, MRF2005544, GNT1149990, fellowships and an investigator grant). We also thank the Victorian Government, especially the Victorian Department of Health and Human Services, the major funder of the Victorian Infectious Diseases Reference Laboratory and Barwon Health. The article reflects the views of the authors and does not represent the views or policies of the funding agencies including the FDA.

### Author Declarations

Institutional ethics committee of the Peter Doherty Institute for Infection and Immunity, Melbourne, Australia (University of Melbourne Central Human Research Ethics Committee gave ethics approval for this work (reference number 2021-21198-15398-3). As this work is part of a prospective cohort study and not an interventional study, registration with ICMJE-approved registry is not required. Human research ethics committee of the Commonwealth Scientific and Industrial Research Organisation (CSIRO) gave ethics approval for this work (reference number 2021_123_RR).

### Summary of Updates

Addressed reviewers' comments and reanalysed with additional Pfizer 3rd dose samples that became available recently. Conclusions unaltered.

